# Do video animations reduce literacy-related inequalities in understanding of health information? A secondary analysis of a systematic review of intervention trials

**DOI:** 10.64898/2026.08.14.26360327

**Authors:** Thirimon Moe-Byrne, Peter Knapp, Su Golder

## Abstract

**Background:** People with lower levels of literacy or health literacy may struggle to understand conventional health information. Video animations show promise as information tools, yet it is unclear whether video animations help reduce these inequalities in understanding. This study examined whether the effectiveness of video animations in health settings differs according to level of literacy or health literacy.

**Methods:** We drew on trials from a recent systematic review of video animations about healthcare or public health topics for patients or the public. We extracted available data on literacy, health literacy, or proxy indicators. One reviewer extracted data and a second checked all entries. Where possible, we conducted subgroup analyses of low and high literacy levels or interaction meta-analyses comparing low versus high literacy groups; otherwise, results were summarised narratively.

**Results:** From 88 eligible trials, we extracted health literacy data for 12. Across nine trials reporting knowledge, animations mostly improved knowledge compared with controls in both lower and higher health literacy groups. Effects on attitudes and behaviours were mixed and often small, with few studies reporting results by health literacy level. Across the subgroup analyses available, there was no consistent evidence of a pooled interaction effect of animations according to low and high literacy groups, but both statistical heterogeneity and small subgroup sizes limited precision of estimates. Across 88 trials, 54 (61%) reported education level, 22 (25%) did not, and 12 (14%) involved children or adolescents likely to have similar education levels.

**Conclusions:** Overall, the available data suggest that video animations can improve knowledge outcomes in both lower and higher health literacy groups, but their impact on attitudes and behaviour is less clear. Because literacy was rarely reported or analysed in the trials, it remains uncertain whether animations help to reduce literacy-related inequalities in access to, and use of health information.

**Author Summary:** Many people find written health information difficult to understand, particularly if they have lower literacy or health literacy. Video animations may make health information easier to follow, but it is not clear whether they help reduce differences in understanding between people with lower and higher literacy levels. We examined 88 trials that tested video animations designed to inform patients or the public about healthcare and public-health topics. We looked for results reported separately for people with different literacy or health literacy levels, or for related measures such as education, socioeconomic deprivation, native language, or cognitive impairment. Only 12 trials provided information that could be used for this purpose.

Overall, animations tended to improve knowledge in both lower and higher literacy groups. However, there was little evidence that they worked better for one group than the other. Effects on attitudes and behaviour were less clear. Most trials did not measure or report literacy-related differences, so we cannot yet tell whether animations reduce, maintain, or widen inequalities in understanding and use of health information. Future studies should include people with lower literacy and report results separately by literacy level.

## Introduction

Video and video animations are increasingly used in healthcare and public health, and have been shown to improve patient knowledge, satisfaction, and information recall. [1] They may also have positive effects on patient satisfaction and health behaviours. [1] Animations may be particularly useful when the information topic is complex or unfamiliar. [2] Animations are a form of multimedia, combining visual and auditory information, and it is claimed that they can make information easier to access and process, particularly for audiences who struggle with conventional text-based materials. [3,4]

Traditionally, health information has been delivered through face-to-face clinical consultations, printed leaflets (with or without images), or short public health broadcasts. However, these methods are not always effective, particularly for individuals with lower levels of literacy or health literacy, who may struggle to process and act on the information provided. [5] Health literacy refers to the ability to obtain, understand, use and critique health information to make appropriate decisions. [6] When these abilities are limited, it can contribute to poorer health outcomes. [7]

While literacy and health literacy (HL) are central to this issue, several proxy indicators can also signal potential challenges in understanding and use of health information. These indicators can include lower levels of formal education, being a non-native language speaker, having an intellectual or learning disability or cognitive impairment, such as dementia, and having a lower socioeconomic background. To address these barriers, video animation, through its combination of visual and auditory elements, offers a promising solution. It can present complex health information in a format that is more engaging and easier to grasp, helping bridge the gap for populations who struggle with traditional communication methods.

However, it is unclear how far existing trials of video animated health interventions have explicitly recruited or reported on their effectiveness in people with low levels of literacy or health literacy or their proxy indicators. In 2026 we published a systematic review of the effectiveness of video animations as information tools across a range of healthcare and public health settings and a variety of population groups; the review included 88 trials.[1] For inclusion in the review, trials had to compare video animations with another form of information provision, such as a booklet or face-to-face consultation, and include at least one of the following outcomes: knowledge, attitudes (such as satisfaction) or behaviour. Based on the 88 trials included in the review, this sub-study has the following aims:

1. To assess how many of the trials exclusively enrolled participants with low levels of literacy or health literacy, or proxy indicators.
2. To assess the effectiveness of video animations in groups with low levels of literacy or health literacy (or proxy indicator) in trials.
3. To assess how many of the trials recruited participants with mixed levels of literacy/health literacy and conducted subgroup analyses, to compare the effects of animation versus control for those with low literacy/health literacy or proxy indicators?
4. To assess the relative effectiveness of video animations in the different literacy or health literacy subgroups in these trials.
5. To assess how many of the trials recruited mixed literacy samples and reported participants’ literacy or health literacy levels (or proxy indicators) in their results, but without undertaking subgroup analysis.

## Methods

This was a secondary analysis of the 88 trials included in a previously published systematic review of video animations as health information tools (see Moe-Byrne 2026). [1] No new database searches or de novo title/abstract screening were undertaken for this sub-study. We systematically re-extracted literacy, health literacy and proxy-indicator data from all reports included in the parent review and conducted additional subgroup and interaction meta-analyses. All 88 trials included in that review were eligible for this sub-study.

### Data extraction and risk of bias

We extracted data on literacy or health literacy variables (or proxy indicators) from all 88 trial reports. One reviewer (TMB) extracted the data, and a second reviewer (PK) checked all entries; disagreements were resolved by consensus. For this sub-study, no additional risk of bias assessment was undertaken; instead, we used the assessment from our published review.

### Data analysis

To examine whether animation effects differed by literacy or health literacy level, we conducted subgroup-specific and interaction meta-analyses. Where subgroup data were available by literacy or health literacy level or a proxy indicator, the Hedges’ g effect size was calculated from the standardised mean difference (SMD) statistic for animation versus control, generated separately in low and high literacy and health literacy subgroups and pooled using a random-effects model. The interaction effect (g interaction) was calculated as the difference in Hedges’ g between subgroups within each trial. A positive g interaction indicates larger animation effects in the low HL group; a negative value indicates larger effects in the high HL group. Statistical significance was set at p < 0.05 with 95% confidence intervals. Heterogeneity was assessed using I² and Q statistics. All analyses were conducted in R (version 4.4.2). Forest plots were produced for each subgroup and for values of g interaction. Where meta-analysis was not possible, data were reported narratively. This secondary analysis is reported in accordance with the PRISMA 2020 statement (see S1 Checklist)

## Results

### Study characteristics

In total, 12 RCTs (from 11 publications) were able to be included in this sub-study (see Fig 1, Table 1). [8,9,3,10–17] Three trials were rated as having low risk of bias, [8,9,16] two as high risk, [11,15] and the remainder as having ‘some concerns’ [3,10,12–14,17] (see Table 1).

**Fig 1.**
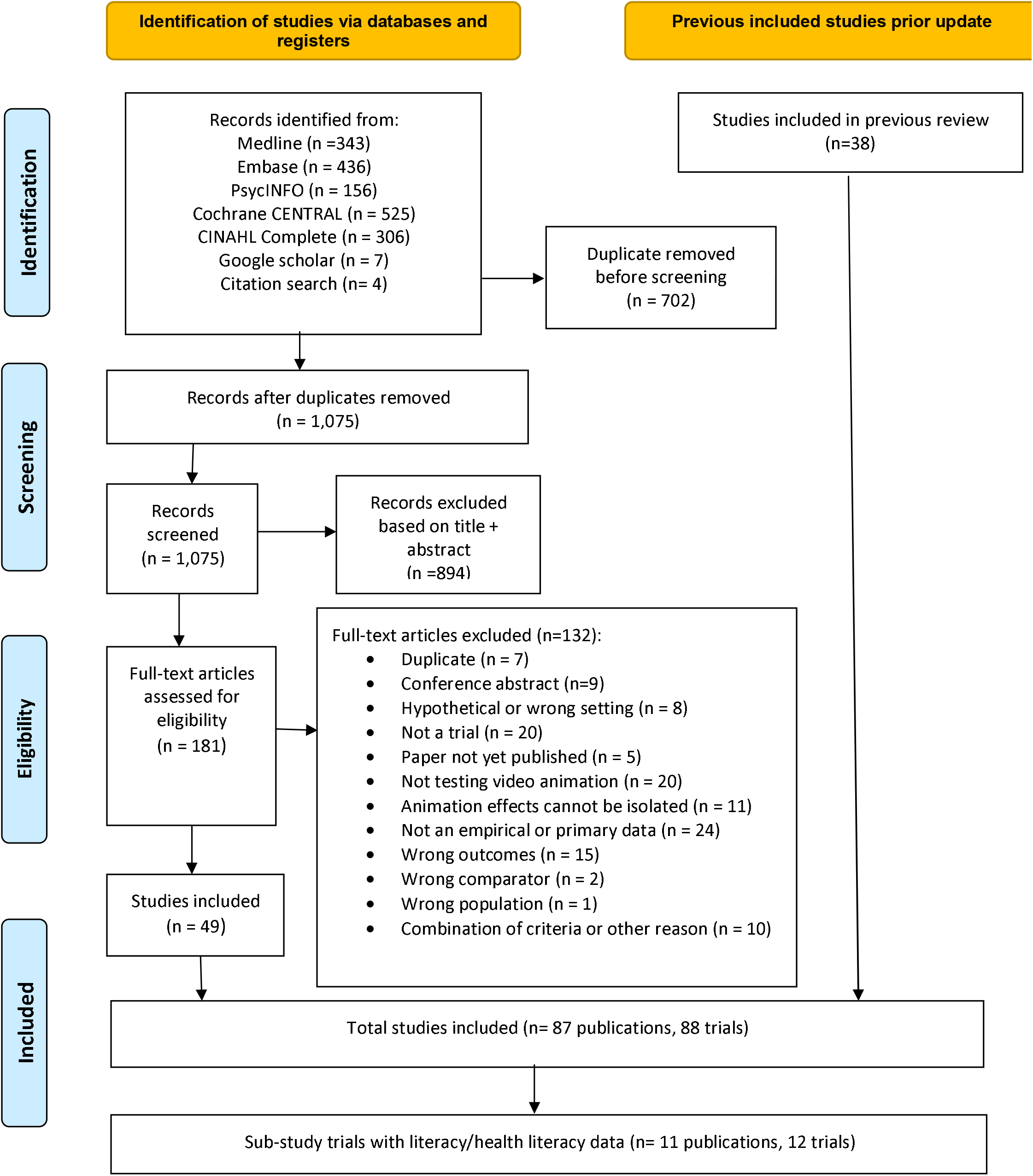
PRISMA Flow Diagram

**Table 1.** Characteristics of the included studies.

### Q1. To assess how many of the trials exclusively enrolled participants with low levels of literacy or health literacy, or proxy indicators

Of the 12 trials only one exclusively enrolled participants with low health literacy or proxy indicators, in this case people with memory impairment [12]. Although another trial [13] included low health literacy in its title, only 19.5% of its participants were classified as having inadequate health literacy.

### Q2. To assess the effectiveness of video animations in low literacy/health literacy groups in these trials

The one eligible trial was a three-arm trial conducted in the Netherlands (n=203) [12] which compared patients at a memory assessment clinic who were undergoing investigative lumbar puncture; they viewed either an educational animation video at home or in the clinic or received usual care (verbal and written information only). Information recall was measured immediately after the consultation (T1), later the same day (T2), and at the end of the day (T3), with significantly higher recall in both of the video animation groups than in the control group at T2 and T3 (T2: F = 23.16, p < 0.001; T3: F = 14.18, p < 0.001). There was no difference in recall at T1. Both video animation groups also reported higher satisfaction with information (means 8.8–8.9 vs 8.4), although this difference was not statistically significant (F = 2.24, p = 0.11).

### Q3. How many trials recruited participants with mixed levels of literacy or health literacy and conducted subgroup analyses, to compare the effects of animation versus control for those with low literacy or low health literacy or proxy indicators?

Ten publications reporting 11 RCTS (two from one publication) recruited participants with mixed levels of literacy or health literacy (or proxy indicators) and conducted subgroup analyses according to levels of literacy or health literacy. Study sample sizes ranged from 79 to 7,803, with a total of 12,403 participants. The trials were conducted in the USA (n = 3), UK (n = 2) and France, Netherlands, Australia, Korea, Canada and Taiwan (one trial each). Two trials used a three-arm design [12,13], and the remaining trials were two-arm [8,9,3,10,11,14–17]. Three trials assessed health literacy using validated instruments such as Short Test of Functional Health Literacy in Adults (STOFHLA) and Short Assessment of Health Literacy in Dutch (SAHL-D), [3,13,15] whereas the remaining trials did not use dedicated health literacy instruments but instead reported proxy indicators such as education level, socioeconomic deprivation and cognitive impairment [8,9,10–12,14,16,17].

### Q4. To assess the relative effectiveness of video animations in low and high literacy and health literacy subgroups

Of the 11 trials that reported subgroup analysis according to literacy or health literacy level, nine assessed knowledge outcomes, [9–11,13,3,8,14,15,17] two examined attitudes and cognition outcomes, [3,11] and two measured behaviour outcomes [16].

#### Knowledge (lower and higher literacy/health literacy group)

Nine studies reported knowledge outcomes; six studies (eight trial arms) [3,8,9,11,13,17] contributed data to meta-analysis; the other three trials could not be included in the meta-analysis as the required data were not included in their findings [10,14,15]. The trial comparators included standard care, static illustrations with spoken or written text, printed text, static images, face-to-face education, or audio booklets. Animation characteristics varied: where reported, durations ranged from 2.16 to 13 minutes, styles included 2D, 3D and cartoon animations, and participant viewing frequency was either once, unlimited over 3 months or not reported.

Video animations were associated with moderate improvements in knowledge for the low literacy/health literacy group (SMD = 0.62, 95% CI 0.13 to 1.10; p = 0.013; I² = 83.4%; 8 RCTs) (Fig 2) and small improvements for the high literacy/health literacy group (SMD = 0.38, 95% CI 0.08 to 0.67; p = 0.012; I² = 70.4%; 8 RCTs) (Fig 3). One study [15] examined Latino or Hispanic patients in the USA with type 2 diabetes across health literacy levels. Participants with low health literacy (STOFHLA < 17) in the intervention group showed statistically significant greater improvements in Diabetes Health Literacy Survey (DHLS) scores compared to controls (61% vs 52%; F = 7.12; p = 0.009), whereas no statistically significant difference was found among those with marginal or adequate health literacy (58% vs 57%; F = 0.82; p = 0.37). A second study [14] found no statistically significant difference in knowledge scores between participants according to their highest education level (school year 12 and below, versus Diploma or higher) (median 6.0 [IQR 4.0–8.0] vs 7.0 [IQR 5.0–9.0]; p = 0.11). The last trial, including kidney transplant candidates [10] evaluated two brief educational animations on kidney donor profile index (KDPI) and increased risk donor (IRD) kidneys, delivered to one group in the trial in addition to standard nurse education, which was provided to the control arm participants. Among older adults with low health literacy, and those with lower educational attainment or lower socioeconomic backgrounds, medium improvements in knowledge were observed in the animation group (Cohen’s d approximately 0.35-0.57). In participants with high health literacy, higher education (college degree or above) or higher income, effects were generally small to medium (Cohen’s d approximately 0.29-0.61).

**Fig 2.**
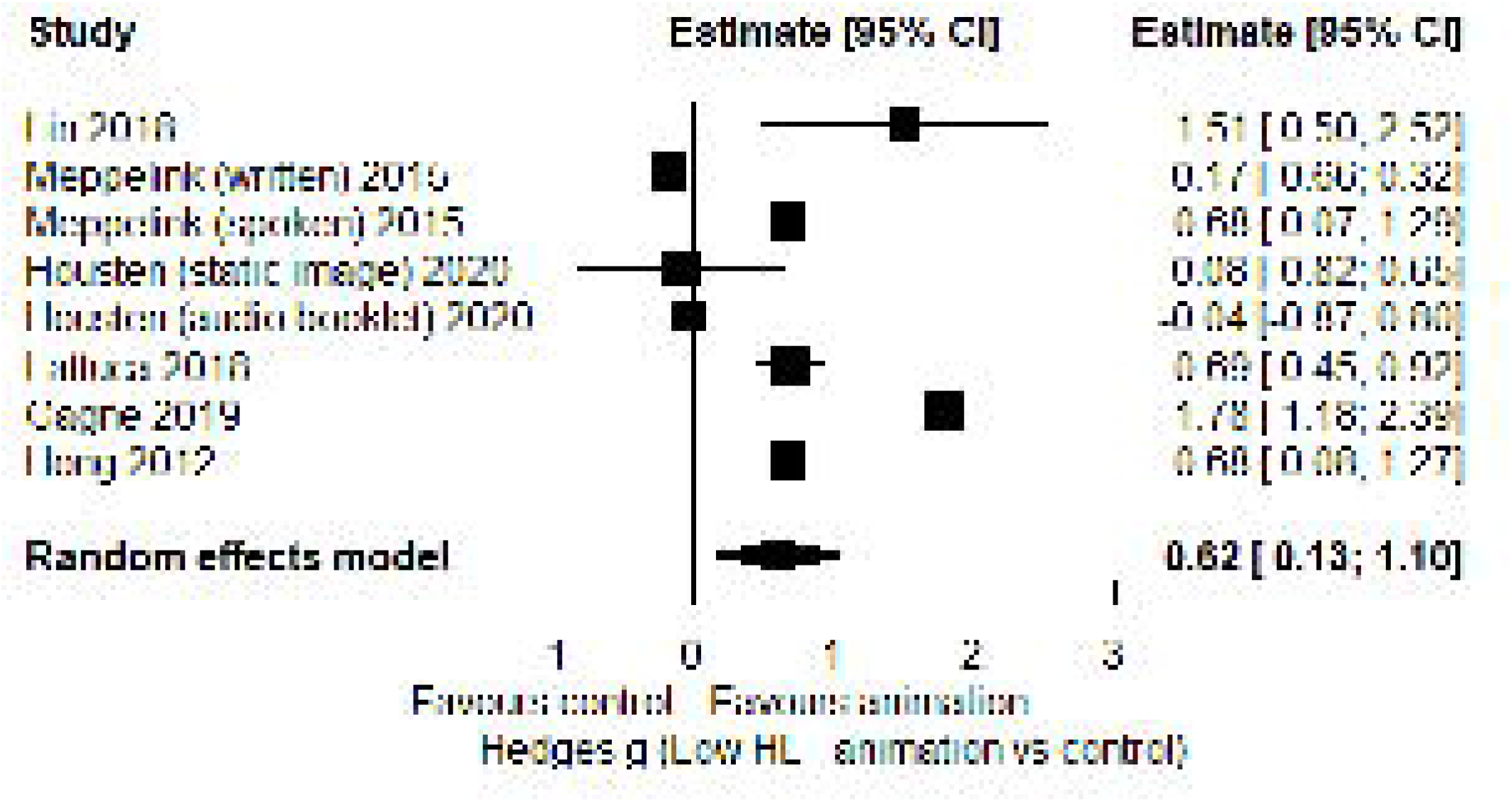
Knowledge outcomes in low literacy/health literacy subgroups (animation vs control), shown as Hedges’ g (standardised mean difference)

**Fig 3.**
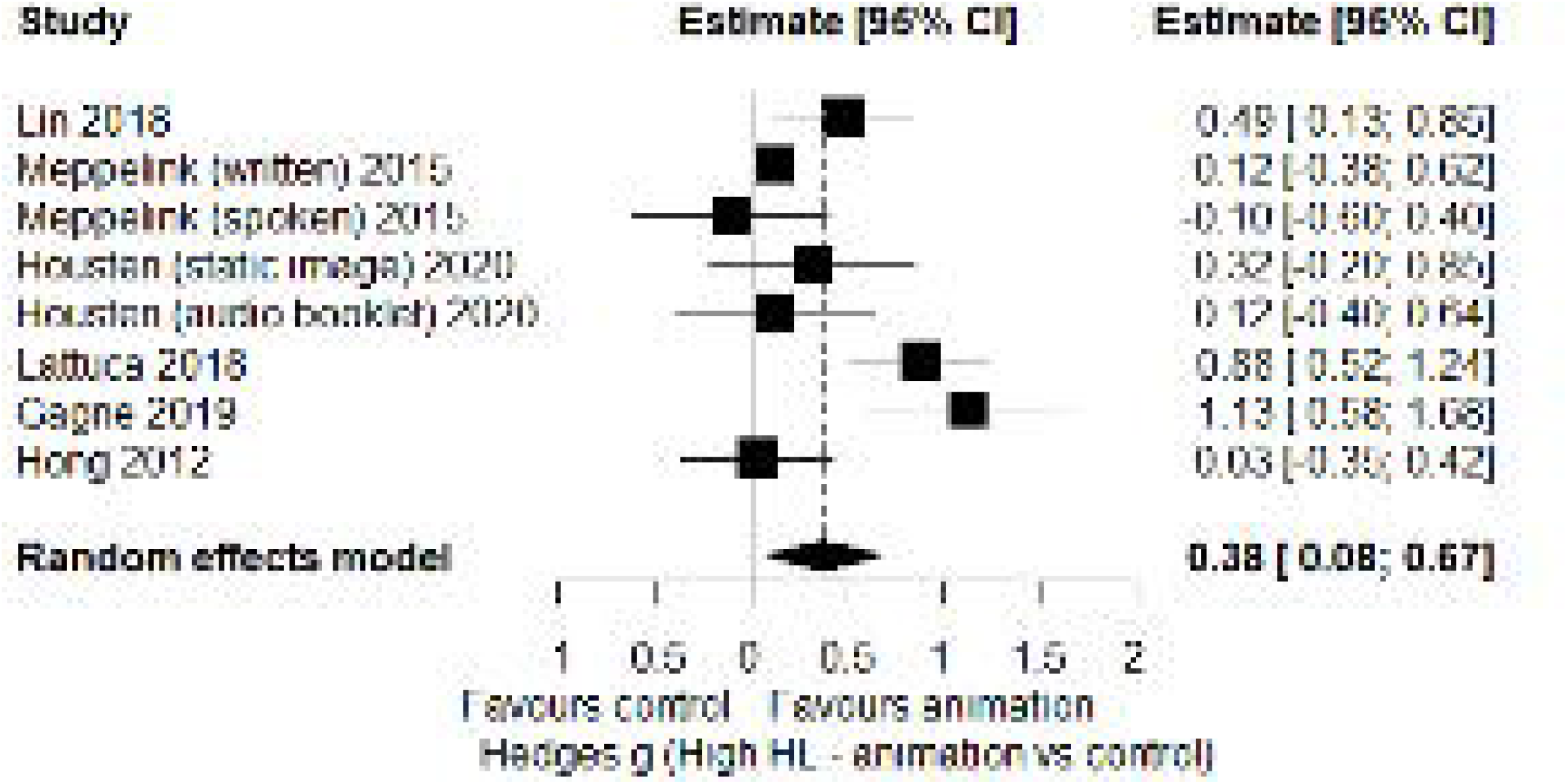
Knowledge outcomes in high literacy/health literacy subgroups (animation vs control), shown as Hedges’ g (standardised mean difference)

#### Knowledge (difference in animation effects between low and high health literacy groups)

Across studies there was no significant interaction between interventions in low and high health literacy groups (pooled interaction g = 0.21, 95% CI [-0.17, 0.59], p = 0.27, I2 = 50.4%; 8 RCTs) (Fig 4), indicating that the effect of animation versus control was similar in participants with different levels of literacy or health literacy.

**Fig 4.**
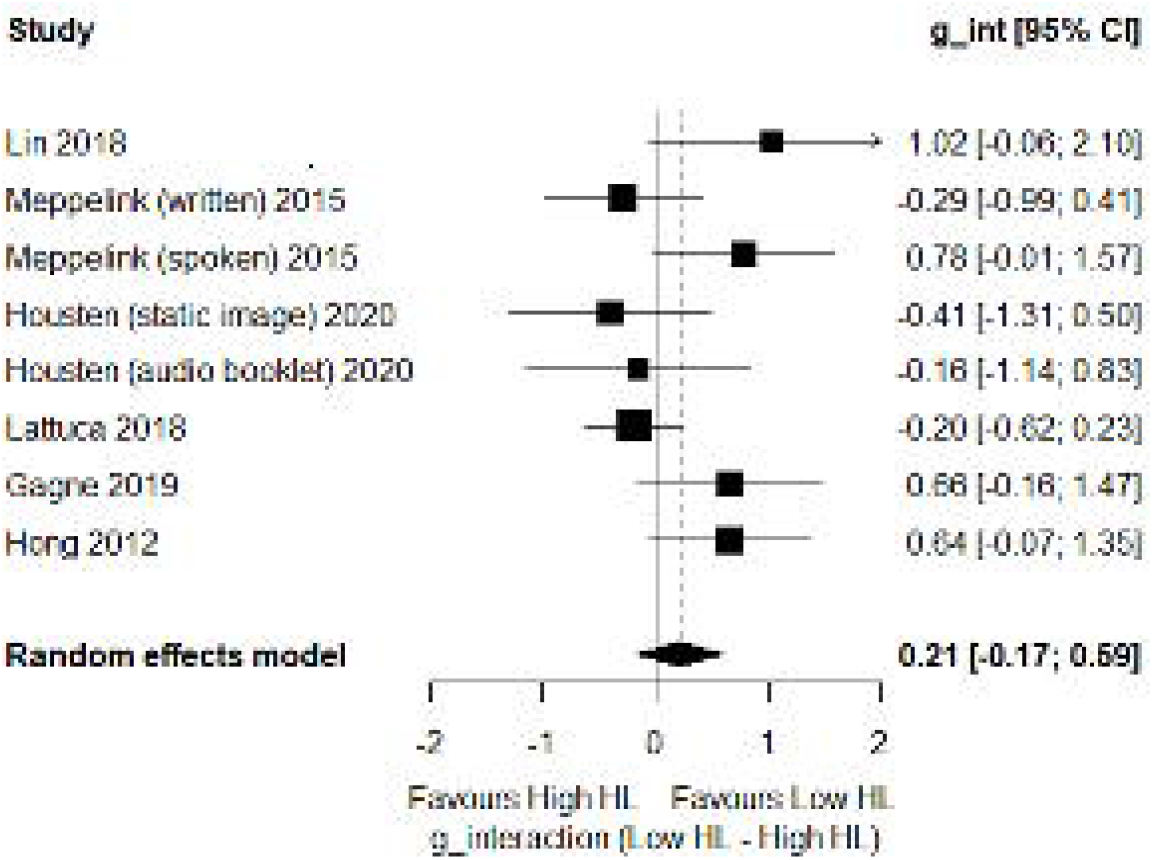
Interaction effect (g interaction) according to low and high literacy/health literacy subgroups

#### Attitude and cognition

Only two trials [3,11] assessed attitude and cognition outcomes according to participants’ health literacy level. One trial [3] delivered colorectal cancer screening messages to older participants (aged ≥55 years) who had either low or high levels of health literacy, comparing video animation with static illustrations. The length and frequency of the animation were not reported. Participants were randomised to receive colorectal cancer screening information as either written (visual text) or spoken (auditory text), combined with either static illustrations or video animations. Thus, there were four conditions: written text with illustrations, written text with animation, spoken text with illustrations, and spoken text with animation. The results showed no statistically significant difference in attitudes toward the message between patients receiving the animation and the static illustration, regardless of text modality (written or spoken) or health literacy level (F = 0.14, p = 0.71).

Another trial [11] enrolled adults who were scheduled to undergo contrast-enhanced CT scan, randomising them to either animation-assisted informed consent or standard verbal informed consent. Participants in the animation arm were asked to watch an animation about the contrast CT scan, lasting 4-5 minutes, shown on a tablet PC. Among patients with less formal education, satisfaction with animation-assisted informed consent was significantly higher in the animation group compared to the control group (8.99 ± 1.13 vs. 7.76 ± 2.31, p = 0.038). No statistical comparison was reported for more highly educated patients.

#### Behaviour

Two RCTs from one publication [16] assessed behaviour outcomes among women aged 50–70 years invited to participate in the UK National Health Service breast screening programme. The two trials differed in their invitation approach: one used timed invitations, in which participants received a fixed, predetermined appointment slot; the other used open invitations, in which participants were offered a choice of appointment times, allowing greater flexibility. Subgroup analyses were conducted among underserved populations, specifically women from high-deprivation areas (IMD deciles 1–5) and those of non-White ethnicity. Both trials compared SMS reminders (informed by behaviour change theory) plus an animation with SMS reminders alone (informed by behaviour change theory). The cartoon animation was 3.5 minutes long with unlimited access to participants over 3 months. A third group in the trial, who received no SMS reminder or animation, was excluded from this analysis as it did not meet our systematic review inclusion criteria for comparing animation with another active information format.

Timed invitation trial: Among participants from more deprived areas who received a timed appointment, attendance was higher in the behavioural SMS plus animation group (83.0%) compared with the behavioural SMS alone group (78.4%), a difference of 4.6 percentage points; however, this difference was not statistically significant (p > 0.05).

Open invitation trial: Among participants from more deprived areas who received an open invitation, attendance rates were similar between the group receiving only a behavioural SMS (53.3%) and the group receiving a behavioural SMS plus an animation (52.1%), with no statistically significant difference observed (p > 0.05).

### Q5. To assess how many trials reported participants’ literacy or health literacy levels (or proxy indicators) in their results, and without undertaking subgroup analysis

Across the 88 trials 54 (61%) reported education level or a proxy measure, of which 12 (14%) provided subgroup data for low and high literacy/health literacy or their proxy and 42 (48%) did not. A further 22 trials (25%) reported no such measures, and 12 (14%) involved children or adolescents likely to have similar education levels.

Seven trials used validated health literacy measures including the Short Assessment of Health Literacy in English (SAHL-E) and Dutch (SAHL-D), [3,18] Short Test of Functional Health Literacy in Adults (S-TOFHLA), [13,15,19] Brief Health Literacy Screener (BHLS), [13] Subjective Numeracy Scale (SNS), [13] Graphical Literacy Measure (GLM), [13] and the Rapid Estimate of Adult Literacy in Medicine–Short Form (REALM-SF) [19–21]. Some trials administered more than one health literacy measure. [13,19]

Ten trials (nine publications) used proxy indicators of socioeconomic status, including the Index of Multiple Deprivation (IMD), [16,22] Scottish Index of Multiple Deprivation (SIMD), [23] household income, [10,11,24] and employment occupation, [19,24–26] but only three [10,11,16] of these ten provided subgroup data for the low literacy or health literacy group when comparing animation versus control.

## Discussion

### Summary of findings

This sub-study analysis of our systematic review [1] examined how video animations have performed for people with low literacy or health literacy, and whether effects differ by literacy or health literacy subgroup.

Across trials that reported subgroups by literacy or health literacy, video animations were associated with moderate improvements in knowledge among participants with lower literacy or health literacy, and smaller but still positive effects among those with higher literacy or health literacy. However, the pooled interaction effect did not indicate a statistically significant difference in animation effectiveness between health literacy groups, implying that animations tend to benefit both groups rather than being of greater advantage to those with lower literacy. However, some individual trials did show patterns consistent with greater gains among low literacy participants highlighting that animations may support greater understanding of information in groups facing greater informational barriers.

Evidence on attitudinal and behavioural outcomes was more limited. Two trials that stratified by health literacy found no clear differences in attitudes or perceptions between animation and comparator formats, and only one reported higher satisfaction among less formally educated participants exposed to animation-assisted consent. Behavioural outcomes, such as breast screening attendance in underserved populations, showed small, non-significant differences when animations were added to behavioural SMS reminders. Together, these findings indicate that animations can enhance knowledge across literacy levels, but their impact on attitudes, satisfaction, and behaviour is less certain.

Despite calls from international and national bodies [27,28] to tackle low health literacy as a contributor to inequalities in health outcomes, only two of the twelve included trials explicitly focused on such participants, in these cases with proxy indicators of low health literacy (memory clinic patients undergoing lumbar puncture and non-native speakers), and very few trials predefined literacy or health literacy groups in their design or analyses. This suggests that most evaluations of health-related animations were conducted in mixed-literacy samples, with little specific attention to those most likely to have difficulty with standard written or spoken information.

### Strengths and Limitations

This review is, to our knowledge, the first to examine the effectiveness of video animations specifically in people with low literacy or low health literacy, using subgroup and interaction meta-analyses where data allowed. It brings together a large body of trial data (88 trials in total), focusing on trials that report literacy or clear proxy indicators, and it applies consistent analytic methods to compare outcomes between lower and higher literacy groups, providing a clearer overview of how animations may work across different literacy levels.

This review also has several limitations. Several trials reported proxy indicators of health literacy but did not present subgroup analyses for these variables. Consequently, relatively few trials contributed to the subgroup and interaction meta-analyses, and there was considerable heterogeneity in populations, settings, comparators and animation formats, as reflected in the high I² values for both low and high literacy groups. Because the subgroup with low health literacy was relatively small, this analysis may have had limited statistical power to detect true differences between low and high literacy or health literacy groups. These factors mean that subgroup effects should be interpreted with caution and they highlight the need for future trials that are large enough to analyse outcomes by literacy level.

### What this study adds

This review shows that, although video animations often improve short-term knowledge, trials rarely report whether their effects differ by health literacy level. Only a small minority of studies directly measured literacy or health literacy with dedicated tools such as STOFHLA or SAHL, while most relied on proxies such as education or lower socioeconomic background. As a result, it remains unclear whether animations make information more accessible for people with low health literacy or whether information benefits are concentrated among those with higher levels of literacy.

### Recommendations for practice

Clinicians and services can use video animations as part of routine information provision, particularly where patients are likely to have low health literacy. However, the evidence on their effects on attitudes, behaviour and longer-term outcomes by literacy level is still limited. Animations should therefore be used alongside other communication approaches, such as plain language and culturally tailored materials, to avoid widening existing inequalities in uptake and use of information.

### Recommendations for Future Research

Although 88 trials were available, we could extract literacy or health literacy outcome data from only 12 trials. This shows that health literacy has received limited attention in trials of health animations. Future research should recruit participants across a wide range of literacy levels, use validated and comparable measures, and pre-specify subgroup analyses by literacy level. Studies should also involve people with low literacy in the co-design of animations, report implementation contexts more clearly, and examine outcomes such as how far animations support shared decision-making in this particular group, whether animations help people in this group stick to treatment, and how animations affect their use of health services. Recent evidence also suggests that the value of health animations depends not only on whether an animation is used, but also on how it is designed, delivered, and tailored to the intended audience. [29] Embedding these literacy sensitive considerations into trial design and analysis will help future research determine more clearly whether animations improve information access and engagement for people who face the greatest barriers.

## Conclusions

Overall, the available data suggest that video animations can improve knowledge in both lower and higher literacy groups. However, their effects on attitudes and behaviour according to health literacy level are less clear, as very few studies have examined these outcomes by literacy level. Across the limited subgroup analyses that were available to undertake, there is no consistent evidence of a pooled interaction effect of animation according to levels of literacy or health literacy. Given the limited data, it remains uncertain whether animations help to reduce literacy-related inequalities in access to understandable health information.

## Supporting information

Table 1. Characteristics of the included studies

## Data availability statement

The original contributions presented in the study are included in the article; further enquiries can be directed to the corresponding author.

## Author contributions

PK and TMB conceived the study. SG conducted the database searches. TMB and PK screened the articles and extracted the data. TMB performed the analysis and drafted the manuscript. All authors contributed to revising the manuscript and approved the submitted version.

