## Supplementary material for "Do video animations reduce literacy-related inequalities in understanding of health information? A secondary analysis of a systematic review of intervention trials": Table 1. Characteristics of the included studies

|  | **Author, year, country** | **Study design** | **Participants, Setting, Education level between the 2 groups** | **Age, Mean (SD) / %Male** | **Total sample;**  **Intervention descriptor & sample size (I);**  **Control descriptor & sample size (C)** | **Intervention details and link (if provided)** | **Results (Intervention vs Control where reported)** (**Knowledge = knowledge or understanding. Attitudes & Cognitions = satisfaction or self-efficacy or confidence in decision, etc. Behaviours = behaviours or skills or intended behaviours) , p value** 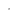 | | | **Risk of Bias** |
| --- | --- | --- | --- | --- | --- | --- | --- | --- | --- | --- |
| 1. | Lin, 2018, Taiwan^9^ | RCT | Adults in Emergency Department due to have acute debridement surgery. | Mean ages not reported.  Male:  I=51%  C=60% | N = 142   < High school  I = 8. Animation about consent + Usual care (verbal information  C= 13  Usual care  ≥ High school  I = 62 Animation about consent + Usual care (verbal information  C= 59  Usual care | 2D graphics animation, including 7 sections about the surgery and aftercare. Included subtitles and captions.  Length not reported.  Link NR. | Knowledge mean (SD):  Post intervention (immediately afterwards):   < High school  I: 46.25(9.16);  C: 35.39(19.42);  P=NS.  ≥ High school  I: 54.84(16.96);  C: 54.24(16.84);  P=NS. | Satisfaction with information. 3 questions, analysed individually. I n=70; C n=72.  Post intervention:  Comprehension: I > C p<0.001.  Helped me with decisions: I > C: p<0.001.  Satisfied with consent process: I > C: p<0.001. | Not assessed. | Low |
| 2. | Kayler 2020, USA^10^ | RCT | Children and adults with range of kidney problems, Erie County Medical Centre (New York). | Age:  I: 61.12(10.39);  C:58.66(10.84)  Male:  I=66.7%;  C:69.4% | N= 79  Health literacy ≤25th percentile  I=10  Animation + audio recorded standard transplant nurse education  C= 9  Audio recorded standard transplant nurse education  Health literacy >25th percentile  I=31  Animation + audio recorded standard transplant nurse education  C= 29  Audio recorded standard transplant nurse education | 2.16 minutes educational video animation about high kidney donor profile index (KDPI) followed by a 2.05-minute animation about increased risk donor (IRD) in addition to audio recorded standard transplant nurse education.  Watched once.  Link:  <https://www.youtube.com/channel/UC3xXkG9VO83Bkj-jS09Vd3Q/videos> | Change in knowledge score mean(SD):  Health literacy ≤25th percentile  I: 2.00 (2.16)  C:1.22 (2.33)  P = 0.461, d= 0.35  Health literacy >25th percentile  I: 2.55 (2.00)  C: 1.41 (1.72)  P=0.022, d = 0.61 | Not assessed. | Not assessed. | Some concerns |
| 3. | Meppelink 2015, Netherlands^3^ | RCT | Colorectal cancer (CRC) screening, participants age 55 years or older with either low or high health literacy. | Age: 68.2 (8.6)  Male:52.4% | N= 231  I= 117  Animation  C= 114  Static illustration  (participants also randomly allocated to receive either spoken or written information) | The animation which included the messages about colorectal cancer screening (i.e. the development of the disease, why early detection is beneficial, the procedure of the test and the possible test outcomes).  Frequency and delivery NR.  Link:  [https://www.ncbi.nlm.nih.gov/pmc/articles/PMC4319081/#app1](https://www.ncbi.nlm.nih.gov/pmc/articles/PMC4319081/%23app1) | Knowledge, mean score out of 28 (95% CI):  Participants with low health literacy (n=108):  Written information:  I: 8.7 (6.9-10.4) (n=35);  C: 9.6 (7.7-11.5) (n=29);  p=NS.  Spoken information:  I: 13.2 (11.0-15.5) (n=21);  C: 9.6 (7.5-11.7) (n=23);  p=0.02.  Participants with high health literacy (n= 123):  Written information:  I: 15.1 (13.2-17.0) (n=29);  C: 14.5 (12.7-16.3)(n=33);  p=NS.  Spoken information:  I: 15.5 (13.7-17.3) (n=32);  C: 16.0 (14.1-17.9) (n=29);  p=NS. | No statistically significant difference in attitudes between animation and static illustration, regardless of text modality or health literacy level (F = 0.14, p = 0.71). | Not assessed. | Some concerns |
| 4. | Calderon 2014, USA^15^ | RCT | Latino/Hispanic patients who were diagnosed with Type 2 diabetes at South Central Family Health Center (SCFHC) and the Charles Drew University of Medicine and Science Center for Health Services Research (Drew). | Age (Range) : 18 to >60  Male: 18.3% | N= 218  Low health literacy  I= 61  Animated video  C= 64 Control (easy-to read text)  Marginal or adequate health literacy  I= 40  Animated video  C= 53 Control (easy-to read text) | 13 minutes animated video which featured an animated icon named “Corazón Quelate” (heart that beats; Spanish version) / “Lotta Hart” (English version). Corazón/Lotta engages viewers with an invitation into her home and emphatically shares her experience with diabetes.  Frequency NR.  Link NR. | Knowledge: Change in DHLS score (STOFHLA scores < 17) mean (%):  Knowledge  Participants with inadequate functional health literacy (I =61, C=52)  61 (53%) vs 52 (50%); F = 7.12, p = 0.009.  Participants with marginal or adequate functional health (I=37, C=50):  37 (58%) and 50 (57%), F = 0.82, p =NS. | Not assessed. | Not assessed. | High |
| 5. | Acharya, 2023(a), UK^16^ | RCT | Women invited for mammography (timed appointments). | Ages not reported by group.  Males: both groups 0%. | N= 2806  I= 1405  Video animation + behavioural text message  C1= 1401  Behavioural text message only | Cartoon animation. Characters of different backgrounds showing the experience of breast cancer screening.  Length 3.5 minutes.  Unlimited access over 3 months.  Link NR. | Not assessed. | Not assessed. | Attendance at screening (undeserved group high IMD decile 1-5, non-White ethnicity):  Timed invitations  I:83% (n=1405);  C1: 78.4% (n=1401)  p=NS. | Low |
| 6. | Acharya, 2023(b), UK^16^ | RCT | Women invited for mammography (open appointments). | Ages not reported by group. | N= 7803  I= 3982  Video animation + behavioural text message  C1= 3821 Behavioural text message only. | Cartoon animation. Characters of different backgrounds showing the experience of breast cancer screening.  Length 3.5 minutes.  Unlimited access over 3 months.  Link NR. | Not assessed. | Not assessed. | Attendance at screening (undeserved group high IMD decile 1-5, non-White ethnicity):  Open invitation  I: 52.1% (n=3982); C1: 53.3% (n=3821);  p=NS. | Low |
| 7. | Housten, 2020, USA^13^ | RCT (3 Arms) | Colorectal cancer screening, participants at a community food bank or the Houston (Texas) Cancer Prevention centre. | Age(range): 45 to 75 years  Male: 36.9% | N= 187  S-TOFHLA adequate  I=41  Video with animated images  C1= 42  Video with static images  C2= 51  Audio booklet  S-TOFHLA inadequate/marginal  I=22  Video with animated images  C1= 20  Video with static images  C2= 11  Audio booklet | 3.3 minutes of the elements of risk communication relevant to colorectal cancer (CRC) screening.  Watched once.  Link provided by the authors (confidential). | Knowledge, total % mean score (SD):  S-TOFHLA adequate  I: 71.6 (15.4);  C1: 65.7 (19.1);  C2: 69.8 (14.5)  P=NS.  S-TOFHLA inadequate/marginal  I: 52.6 (18.0);  C1: 54.3 (21.2);  C2: 53.3 (18.2)  P=NS. | Not assessed. | Not assessed. | Some concerns |
| 8. | Gagne, 2019, Canada^17^ | RCT | Adults with atrial fibrillation, hospital department in Quebec. | Age mean(SD) :  I: 57 (13); C: 56 (13)  Males: I: 77%; C: 60%. | N= 60  I= 30  Video animation + face to face education session from nurse (45 minutes)  C= 30  Face to face education session from nurse (45 minutes) | Video on AF, including normal heartbeat, symptoms of AF, risk factors for AF, complications of AF, and treatment options (ie same topics as in the f2f session).  Length: 8 minutes.  Suggests video watched once only.  Link NR. | Knowledge: knowledge of AF test (KAF), focussed on symptoms, disease, impact, risk of stroke. 25 items. Range 0-25 (higher is better). Immediately after:  ≤High school diploma/Vocational studies diploma (n=27)  I: 19.8(1)  C:17.9(1.1)  P=0.0048.  ≥Postsecondary diploma below bachelor level/University diploma (n=33)  I: 20.9 (0.7)  C: 20.1 (0.7)  P=0.52 | Not assessed. | Not assessed. | Some concerns |
| 9. | Hong, 2012, Korea^11^ | RCT | Emergency department patients about to undergo CT scan | Age: I= 38 (13.7); C= 42.3 (14.0),  Male:  I= 45%;  C= 56% | N= 150  I= 75  Animation + Usual care (verbal informed consent)  C= 75  Usual care | 7 minutes animation assisted informed consent about the contrast Computed Tomography (CT) scan.  Watched once on tablet computer.  Link NR. | Knowledge, combined mean score (SD).  Post intervention:  ≤High school (n=47)  I: 8.73 (0.27)  C: 7.53(2.28)  ≥ University (n= 103)  I: 8.57 (1.50)  C: 8.52 (1.40) | Satisfaction, combined mean (SD).  Post intervention:  ≤High school (n= 47)  I: 8.99 (1.13)  C: 7.76(2.31)  ≥ University (n= 103)  I: 8.67 (1.39)  C: 8.29 (1.67) | Not assessed. | High |
| 10. | Gois, 2024, Australia^14^ | RCT | Patients undergoing clinically indicated percutaneous kidney biopsies | Age: Median(range):  52 (IQR 34-65)  Male: 44% | N=120  I= 60  Video + Usual care (verbal information to inform consent)  C= 60  Usual care (verbal information to inform consent) | 8 minutes explanatory animation on an online platform  covering the procedure, its risks, and pre- and post-biopsy care before providing digital consent. The video allowed  participants to pause or rewind as needed.  Link NR. | Knowledge (possible range 0-9):  Median (IQR): Post-intervention  Year 12 and below (n=70): median 6.0, IQR 4.0–8.0  Diploma or higher (n=49): median 7.0, IQR 5.0–9.0 | Not assessed. | Not assessed. | Some concerns |
| 11. | Lattuca 2018, France^8^ | RCT | Patients undergoing coronary angiography and/or angioplasty,  Cardiology units (39 participating centres in France) | Age: NA  Male: NA | N= 425  I= 207  3D educational video + Usual care  C= 218  Usual care | 5 minutes 3D education video on the procedure of the coronary angiography  Watched once on a tablet.  Link:  <https://ars.els-cdn.com/content/image/1-s2.0-S0002870318300784-mmc1.mp4> | Knowledge, mean total score out of 16 (SD):  Post intervention:  Primary school level(n=293):  I: 10.7(3.2);  C: 8.5(3.2);  P<0.001  University level education(n=132):  I: 13.2(2.1);  C11.1(2.6)  P<0.001 | Not assessed for subgroups | Not assessed. | Low |
| 12. | Mofrad 2021, Netherlands^12^ | RCT (3 arms) | Patients visiting the memory clinic of the Alzheimer Centre Amsterdam. | Age: 63 (9)    Male:  I1: 53%; I2: 69%; C: 70% | N= 209  I1= 63  Animation viewing at home + Usual care (verbal information and an informational folder on the LP procedure)  I2 =70  Animation viewing in clinic + Usual care  C= 76  Usual care | 3 minutes animation video to inform and prepare patients and caregivers for the Lumbar puncture (LP) procedure in the context of Alzheimer’s Disease (AD) diagnosis which included the LP procedure and the most common complications.  All participants also received care as usual.  Home viewing group were allowed to watch the video as often as desired. Clinic viewing group viewed the video once in the waiting room.  Link NR. | Information recall:  T2 (screening day)  I1 vs C:  mean difference 0.97 (0.29) (I1 n=62; C n=74) (p=0.003).  I2 vs C:  mean difference 1.35 (0.28) (I2 n=67; C n=74) (p<0.001).  T3 (end of screening day)  I1 vs C:  mean difference 0.84 (0.26) (I1 n=62; C n=74); (p=0.005).  I2 vs C:  mean difference 0.9 (0.26); (I2 n=67; C n=74); (p=0.002).  There was no difference in information recall between home and clinic viewing at either T2 or T3. | Satisfaction: no difference in satisfaction between interventions and control in terms of satisfactory F [2,176] = 2.24,  (p=NS).  (I1 n=61; I2 n=67; C n=74) | Not assessed. | Some concerns |
